# Pressure is not flow: Rethinking blood pressure as a surrogate for cerebral blood flow during general anesthesia

**DOI:** 10.64898/2026.10.07.26365014

**Authors:** Jonas Österlind, Petter Holmlund, Johan Birnefeld, Magnus Hultin, Anders Wåhlin, Laleh Zarrinkoob

## Abstract

**Background:** Mean arterial pressure (MAP) is pharmacologically augmented in selected perioperative and critical care settings, including acute neurological conditions and after cardiac arrest, with the aim of improving cerebral perfusion, although it remains uncertain whether higher arterial pressure translates into increased cerebral blood flow (CBF). We investigated the CBF response to MAP augmentation with norepinephrine (NE), a commonly used vasopressor in perioperative and critical care settings, during propofol-based general anesthesia.

**Methods:** In this prospective within-subject interventional study, 16 neurologically intact patients with ASA physical status I–II undergoing elective surgery were studied using phase-contrast magnetic resonance imaging. CBF was quantified in the awake state and during propofol-based general anesthesia before and after NE-induced MAP augmentation. During general anesthesia, NE was titrated to increase MAP by approximately 25% from baseline. Concurrent systemic hemodynamics were assessed.

**Results:** General anesthesia was associated with a 50.1% (interquartile range, 42.1–55.7%) reduction in CBF and a 24.2% (20.3–29.2%) reduction in MAP compared with the awake state (both P<0.001). During general anesthesia, NE-induced MAP augmentation increased MAP by 25.7% (21.9–32.0%; P<0.001), whereas CBF remained unchanged (P=1.000). Cardiac output remained unchanged (P=0.163), whereas systemic vascular resistance index and cerebrovascular resistance increased by 40.0% (28.7–45.2%) and 36.2% (17.7–42.8%), respectively (both P<0.001). Changes in CBF during MAP augmentation were not associated with concurrent changes in MAP, cardiac output, or systemic vascular resistance index.

**Conclusions:** In neurologically intact patients undergoing propofol-based general anesthesia, clinically relevant NE-induced MAP augmentation did not increase CBF despite an approximately 50% lower CBF than in the awake state. These findings challenge the assumption that pharmacological elevation of an otherwise adequate MAP improves cerebral perfusion in this setting. While maintaining adequate arterial pressure remains essential, further MAP augmentation should not be assumed to increase CBF.

Clinical Trial Registration: https://clinicaltrials.gov/study/NCT06855407; NCT06855407.

**Clinical Perspective:** *What Is New?:*

- Norepinephrine-induced augmentation of mean arterial pressure by approximately 26% during propofol-based general anesthesia did not increase cerebral blood flow in neurologically intact patients.
- Cerebral blood flow was approximately 50% lower during general anesthesia than in the awake state and remained at this lower level during mean arterial pressure augmentation.
- Cardiac output remained unchanged, whereas systemic and cerebrovascular resistance increased during norepinephrine-induced mean arterial pressure augmentation.

*What Are the Clinical Implications?:*

- Pharmacological augmentation of an otherwise adequate mean arterial pressure should not be assumed to increase cerebral blood flow during propofol-based general anesthesia.
- Mean arterial pressure alone may therefore be an inadequate surrogate for cerebral blood flow when setting hemodynamic targets during general anesthesia.
- Maintaining adequate arterial pressure remains essential, but further pharmacological pressure augmentation should be distinguished from the treatment of hypotension.

## Introduction

Mean arterial pressure (MAP) augmentation is used or investigated across a range of acute neurological conditions, following cardiac arrest, and in perioperative care, with the aim of maintaining or increasing cerebral perfusion and ultimately improving neurological outcomes.^1–4^ However, evidence of clinical benefit has thus far been limited. Although the arterial pressure required to maintain adequate cerebral perfusion is likely patient-specific,^5^ MAP augmentation can be conceptually distinguished from treatment of hypotension by the intentional elevation of arterial pressure to a higher target rather than merely correcting a low one.^6^ Many of these clinical contexts involve sedation or general anesthesia, during which cerebrovascular tone is altered and cerebral perfusion, as reflected by cerebral blood flow (CBF), may decrease in parallel with reduced cerebral metabolic demand.^7,8^ The relationship between MAP and CBF under these conditions therefore warrants investigation.^9^

Norepinephrine (NE) is commonly used for blood pressure management during general anesthesia^10^ and is recommended as the first-line vasopressor for acute circulatory failure in international critical care guidelines.^11,12^ Although NE reliably increases arterial pressure primarily through systemic α1-mediated vasoconstriction, it may also exert direct cerebrovascular effects.^13^ Reductions in cerebral oxygenation and flow velocity despite increased MAP have been described,^14^ and meta-analytic data indicate that changes in cerebral blood flow velocity during pharmacological MAP augmentation depend on autoregulatory status.^15^ However, blood flow velocity provides only a surrogate measure of CBF rather than direct quantification.^16^ Phase-contrast magnetic resonance imaging (PC-MRI) enables noninvasive measurement of blood flow in large vessels and quantitative assessment of CBF.^17^ In healthy volunteers studied in the awake state, we previously demonstrated a reduction in CBF during NE-induced MAP augmentation.^18^

Despite the clinical use of pharmacological MAP augmentation, its effect on CBF during general anesthesia has not been quantitatively determined. Using PC-MRI, we aimed to determine the CBF response to NE-induced MAP augmentation during propofol-based general anesthesia. Measurements in the awake state served as a physiological reference, and concurrent systemic hemodynamics were evaluated.

## Methods

### Study Design

This prospective, within-subject interventional study investigated the effect of NE-induced MAP augmentation on CBF during general anesthesia. Participants underwent PC-MRI in the awake state and at two time points during general anesthesia, before and after NE-induced MAP augmentation, with each participant serving as their own control. The study was conducted at Umeå University Hospital, Sweden, according to a prospectively published study protocol,^19^ portions of which are included here to ensure transparency and reproducibility. The study was registered at ClinicalTrials.gov (NCT06855407), approved by the Swedish Ethical Review Authority (2024-00251-01; amendment 2024-05508-02), conducted in accordance with the Declaration of Helsinki, and all participants provided written informed consent. One author (J.Ö.) had full access to all study data and takes responsibility for its integrity and analysis.

### Participants

Adults aged 50 to 80 years scheduled for elective otolaryngological surgery under general anesthesia were consecutively recruited between January 2025 and April 2026. Eligible participants had a Mini-Mental State Examination (MMSE) score >23 and American Society of Anesthesiologists (ASA) physical status I–III. Patients with a body mass index (BMI) >30 kg/m², intracranial pathology, previous major cerebrovascular disease, contraindications to MRI, or contraindications to NE-induced MAP augmentation were excluded.

### Study Procedures

Detailed study procedures have been reported in the published study protocol.^19^ Briefly, the awake MRI examination was performed on the day before surgery. On the day of surgery, participants underwent the planned otolaryngological procedure under general anesthesia, followed by MRI. Anesthesia during MRI was maintained with a continuous infusion of propofol (4–8 mg·kg⁻¹·h⁻¹), a commonly used intravenous hypnotic agent for general anesthesia and sedation in critical care.^20–22^ The propofol infusion rate was kept constant throughout both the baseline and augmented MAP measurements. A fentanyl bolus (150–200 µg) was administered before MRI. Mechanical ventilation was adjusted to maintain stable end-tidal carbon dioxide throughout the protocol. During baseline MRI under general anesthesia, NE was administered as required to maintain MAP at the same clinically accepted level as during the preceding surgical procedure, with a minimum MAP of 65 mmHg. NE was subsequently titrated stepwise every 2 minutes by 0.01–0.04 µg·kg⁻¹·min⁻¹ to achieve the augmented MAP state, defined as an approximately 25% increase in MAP relative to baseline, with a safety limit of systolic arterial pressure ≤160 mmHg. MRI acquisition at both baseline and augmented MAP was performed after a hemodynamic steady state had been achieved, defined as a MAP variation ≤3 mmHg for approximately 5 minutes. During general anesthesia, invasive arterial pressure, heart rate, oxygen saturation, and end-tidal carbon dioxide were continuously monitored. Awake blood pressure was measured noninvasively immediately before induction of anesthesia.

All MRI examinations were performed on a 3.0-T clinical MRI system. Three-dimensional T1-weighted images were acquired for brain volume estimation, and CBF was quantified using PC-MRI.^23^ Imaging planes were positioned perpendicular to the internal carotid and vertebral arteries at the C2–C3 level and the ascending aorta for assessment of systemic flow. Flow quantification was performed using Segment version 3.2 R9074 (Medviso AB, Lund, Sweden)^24^ with semiautomatic vessel segmentation when feasible and manual ROI placement when required. All datasets were analyzed by a single observer (J.Ö.). Interobserver reproducibility was assessed in five randomly selected participants by having a second observer (P.H.) independently repeat ROI placement and flow quantification of the bilateral internal carotid and vertebral arteries (60 vessel measurements). Intraclass correlation coefficients were calculated using a two-way mixed-effects model for absolute agreement and single measurements (ICC[3,1]).^25^ Additional details regarding physiological monitoring and MRI acquisition are provided in the Supplemental Methods.

CBF was calculated as the sum of blood flow in the bilateral internal carotid and vertebral arteries. Cardiac output was derived from ascending aortic flow. Cardiac index was calculated as cardiac output divided by body surface area, stroke volume as cardiac output divided by heart rate, and stroke volume index as stroke volume divided by body surface area. Cerebrovascular resistance was calculated as (MAP − intracranial pressure)/CBF, assuming an intracranial pressure of 10 mmHg,^26^ and systemic vascular resistance index as ([MAP − central venous pressure]/cardiac index) × 80, assuming a central venous pressure of 10 mmHg.^27^

### Outcome Measures

The primary outcome was the change in CBF between baseline and the augmented MAP state during general anesthesia. Measurements obtained in the awake state served as a physiological reference. Secondary outcomes included changes in systemic hemodynamic variables and exploratory associations between changes in CBF and concurrent changes in MAP, cardiac output, and systemic vascular resistance index, both from the awake state to general anesthesia and during NE-induced MAP augmentation.

### Statistical Analysis

Continuous variables are reported as median (interquartile range). Analyses were performed on complete paired datasets. Given the within-subject design and limited sample size, nonparametric statistical methods were used throughout.^28^ Paired comparisons between physiological conditions were performed using the Wilcoxon signed-rank test. Associations between relative changes in CBF and concurrent relative changes in MAP, cardiac output, and systemic vascular resistance index from the awake state to general anesthesia and during NE-induced MAP augmentation were explored using Spearman’s rank correlation. Both sexes were included in the study. Given the limited sample size, analyses were performed on pooled data and were not stratified by sex. Secondary analyses were exploratory, were not adjusted for multiple comparisons, and should be interpreted as hypothesis-generating. For comparison with conventionally reported tissue-normalized CBF values, CBF was additionally normalized to MRI-derived brain volume and expressed per 100 g of brain tissue, assuming a tissue density of 1.03 g/mL.^29^ All statistical tests were two-sided, and a P value <0.05 was considered statistically significant. Statistical analyses were performed using SPSS version 32 (IBM Corp., Armonk, NY, USA).

A clinically meaningful increase in CBF was prespecified as ≥15% relative to baseline. This threshold exceeds expected measurement repeatability and short-term biological variability for PC-MRI^30^ and was considered conservative relative to previous physiological studies.^15^ Sample size was calculated to provide 95% power with a two-sided α of 0.05 to detect a ≥15% increase in CBF associated with NE-induced MAP augmentation. The calculation was based on the standard deviation of within-subject CBF change reported in a previous experimental study,^18^ which was conservatively doubled to account for the greater expected physiological heterogeneity during general anesthesia.

## Results

Eighteen participants were enrolled. One participant was excluded before any study procedures, as baseline blood pressure was elevated and protocol-driven MAP augmentation was considered inappropriate by the attending anesthesiologist. One participant experienced neck discomfort during the MRI examination in the awake state, resulting in premature termination of the examination, and was subsequently not studied under general anesthesia. Sixteen participants completed the study and were included in the final analysis. Baseline characteristics of the study population are presented in **Table 1**. Interobserver reproducibility for the flow analysis was excellent (ICC[3,1] = 0.991; 95% CI, 0.975–0.997).

**Table 1.** Baseline Characteristics of the Study Population.

| Variable | Participants (n=16) |
| --- | --- |
| **Demographics** |  |
| Age, years | 63 (59–71) |
| Female sex, n (%) | 6 (38) |
| Height, cm | 173 (161–178) |
| Weight, kg | 77 (70–85) |
| BMI, kg/m<sup>2</sup> | 26.6 (23.9–28.3) |
| **Clinical characteristics** |  |
| Chronic hypertension, n (%) | 8 (50) |
| Lung disease, n (%) | 4 (25) |
| Nicotine use, n (%) | 4 (25) |
| **ASA physical status, n (%)** |  |
| I | 2 (12.5) |
| II | 14 (87.5) |

Data are presented as median (IQR) or number (%). BMI indicates body mass index; ASA, American Society of
Anesthesiologists.

Induction of general anesthesia was associated with a 50.1% (42.1–55.7%; P<0.001) reduction in CBF and a 24.2% (20.3–29.2%; P<0.001) reduction in MAP compared with the awake state (Table 2; Figure 1). Cardiac output decreased by 14.3% (1.3–28.4%; P=0.015). During general anesthesia, titration of NE infusion from 0.035 (0.010–0.050) to 0.095 (0.080– 0.150) µg·kg⁻¹·min⁻¹ increased MAP by 25.7% (21.9–32.0%; P<0.001), achieving the predefined target increase. CBF remained unchanged (**Figure 1**; P=1.000). Cardiac output remained unchanged (P=0.163), whereas systemic vascular resistance index and cerebrovascular resistance increased by 40.0% (28.7–45.2%; P<0.001) and 36.2% (17.7–42.8%; P<0.001), respectively (**Table 2**).

**Table 2.**
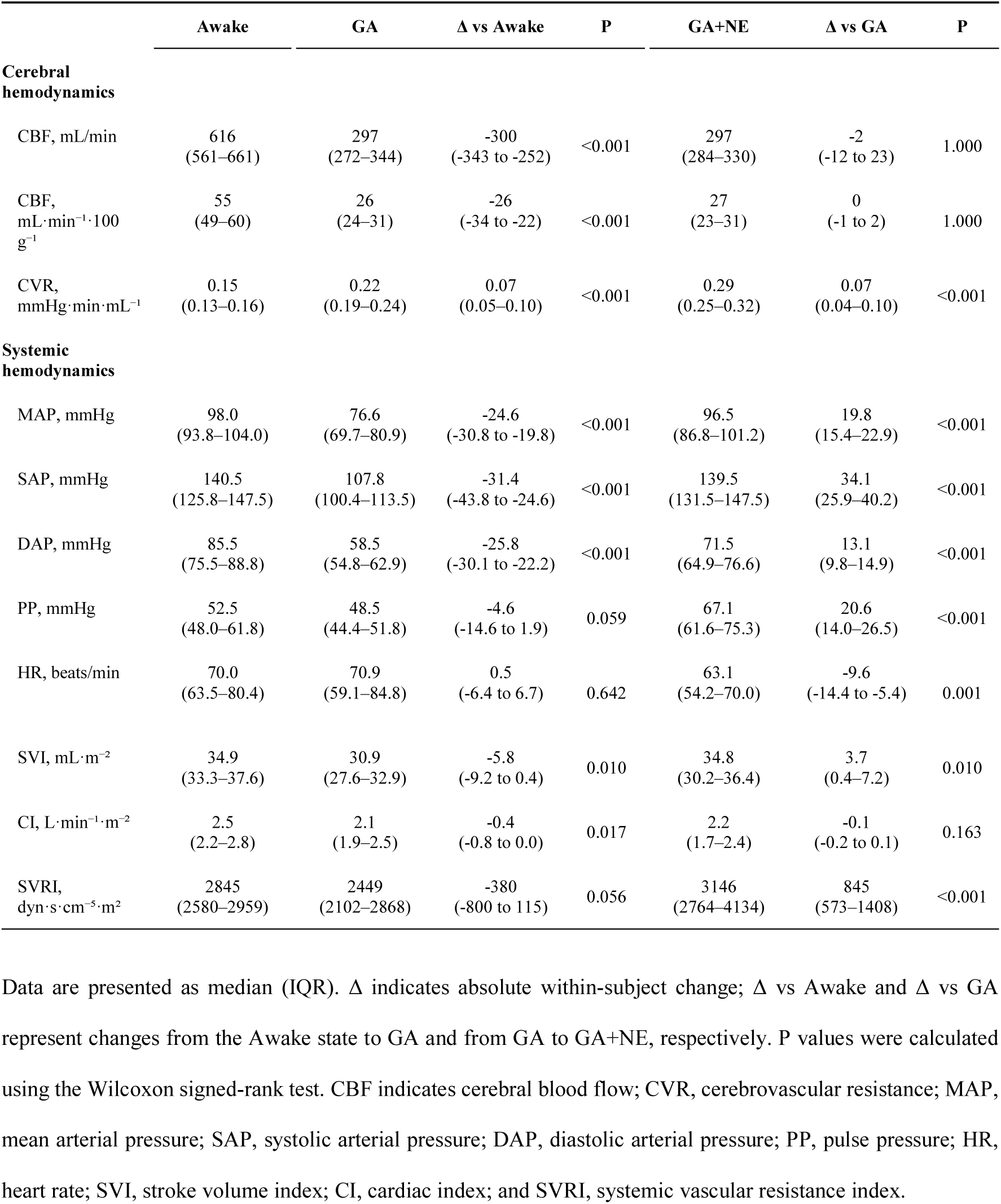
Cerebral and systemic hemodynamic variables in the awake state, general anesthesia, and norepinephrine-induced mean arterial pressure augmentation.

**Figure 1.**
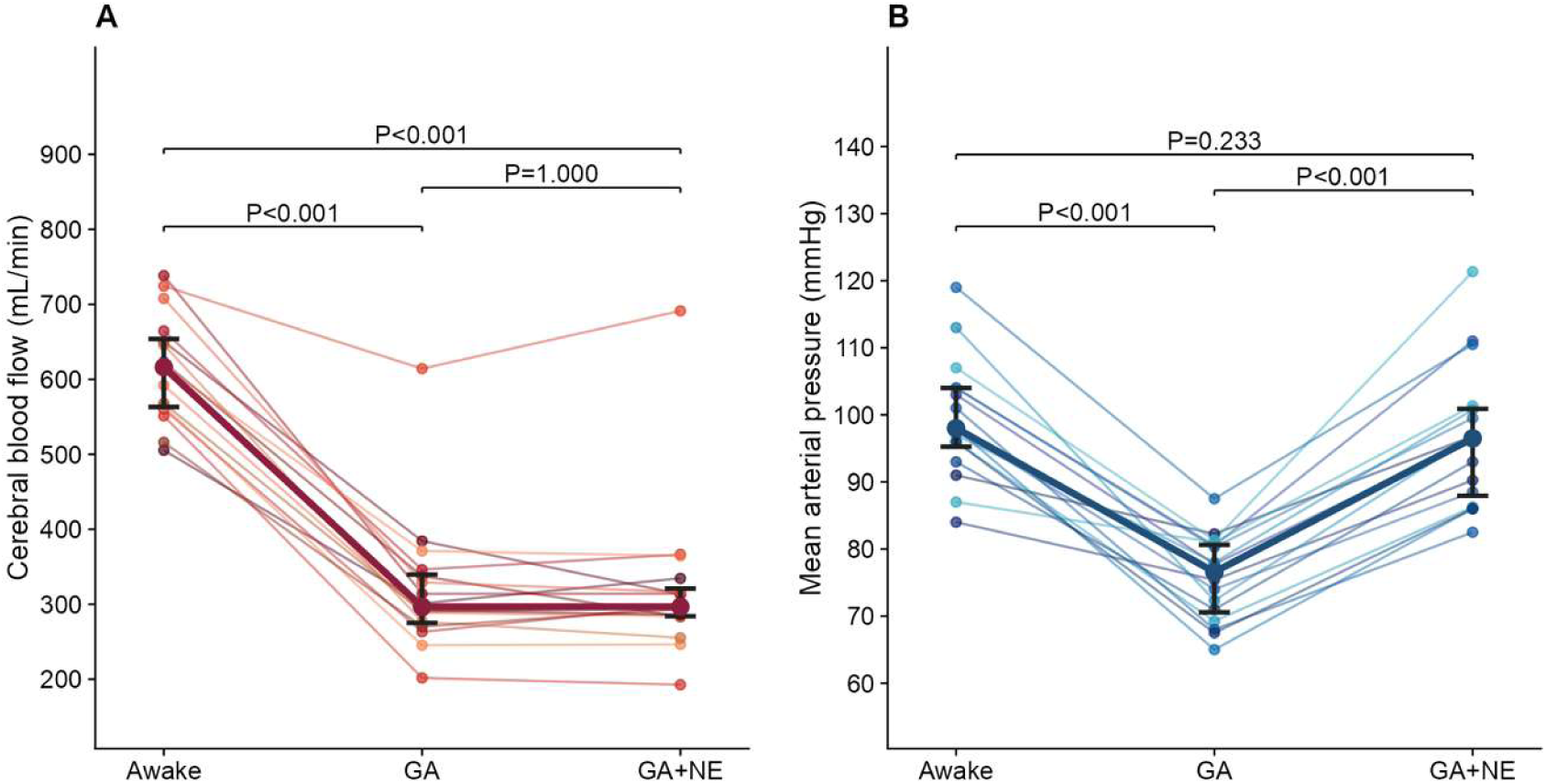
Effect of norepinephrine-induced mean arterial pressure augmentation on cerebral blood flow during general anesthesia. A, Cerebral blood flow (CBF). B, Mean arterial pressure (MAP). Measurements were obtained in 16 participants in the awake state (Awake), under general anesthesia (GA), and after norepinephrine titration during general anesthesia (GA+NE). Norepinephrine was titrated to achieve the predefined target increase in MAP. Thin lines represent individual participants, whereas thick lines and error bars indicate the median and interquartile range. P values are shown for pairwise comparisons using the Wilcoxon signed-rank test.

Exploratory analyses of relative changes from the awake state to general anesthesia showed a moderate correlation between changes in CBF and cardiac output (ρ=0.55, P=0.026), whereas no statistically significant correlations were observed with changes in MAP (ρ=−0.11, P=0.68) or systemic vascular resistance index (ρ=−0.48, P=0.058). During NE-induced MAP augmentation, no significant associations were observed between relative changes in CBF and concurrent changes in MAP (ρ=−0.19, P=0.47), cardiac output (ρ=0.08, P=0.78), or systemic vascular resistance index (ρ=−0.35, P=0.19).

Ventilator settings remained unchanged throughout the MRI study protocol, with end-tidal carbon dioxide maintained at 4.8 (4.7–5.0) kPa.

## Discussion

MAP augmentation is currently used in clinical practice on the premise that higher arterial pressure maintains or increases cerebral perfusion. The present study provides novel physiological findings that challenge this concept. First, NE-induced MAP augmentation during propofol-based general anesthesia did not increase CBF despite a clinically relevant 26% increase in MAP. Second, CBF during general anesthesia was approximately 50% lower than in the awake state and remained at this lower level during MAP augmentation.

The absence of an increase in CBF during MAP augmentation is consistent with cerebral autoregulation. With stable intracranial pressure, increases in MAP translate into increased cerebral perfusion pressure (CPP). Within the autoregulatory range, increases in CPP are counteracted by changes in cerebrovascular tone, thereby limiting pressure-dependent changes in CBF. In the present study, CBF remained unchanged despite increased MAP and consequently increased CPP. Although autoregulation was not directly assessed, this pressure– flow response is consistent with preserved autoregulatory capacity and previous observations of minimal changes in cerebral blood flow velocity during vasopressor-induced MAP augmentation under intravenous anesthesia.^31^

The systemic hemodynamic response to NE was characterized by a marked increase in systemic vascular resistance, whereas cardiac output remained unchanged, illustrating the fundamental distinction between arterial pressure and blood flow. Changes in CBF were not associated with concurrent changes in MAP, cardiac output, or systemic vascular resistance index, providing no evidence that variability in the systemic hemodynamic response to NE explained the interindividual differences in CBF during MAP augmentation.

These physiological findings provide context for randomized trials evaluating different blood pressure targets in critically ill patients. In patients with septic shock, targeting a MAP of 80–85 mmHg rather than 65–70 mmHg did not improve survival in the SEPSISPAM trial.^32^ Similarly, randomized trials evaluating higher MAP targets after cardiac arrest have not demonstrated improvements in neurological recovery or survival,^33,34^ findings also supported by meta-analysis.^35^ In this context, recent evidence after endovascular treatment for acute ischemic stroke suggests that blood pressure management tailored to the cerebral pathophysiological context may improve functional outcome compared with a uniform blood pressure target.^36^ Although these studies differ substantially in population, intervention, and clinical end points, collectively they highlight the limitations of applying uniform blood pressure targets across heterogeneous physiological states. Our findings complement this perspective by demonstrating that, during general anesthesia, further augmentation of an otherwise adequate MAP does not necessarily increase CBF.

Any potential benefit of higher MAP targets must also be weighed against the risks associated with greater vasopressor exposure. In the SEPSISPAM trial, the higher MAP target was associated with an increased incidence of new-onset atrial fibrillation.^32^ Similarly, the randomized HIMALAIA trial of induced hypertension for delayed cerebral ischemia after aneurysmal subarachnoid hemorrhage was associated with a numerically higher rate of serious adverse events, although the trial was terminated early because of slow recruitment and the absence of an observed effect on cerebral perfusion.^37^ Together with the present findings, these observations emphasize the distinction between correcting hypotension to maintain adequate perfusion pressure and further pharmacological augmentation of MAP once an adequate arterial pressure has been achieved.

General anesthesia was associated with an approximately 50% reduction in CBF compared with the awake state. This change was moderately correlated with the concomitant reduction in cardiac output. However, the relative decrease in CBF was substantially greater than the 14% reduction in cardiac output, suggesting that reduced systemic flow alone does not explain its magnitude. Rather, reduced cerebral metabolism during propofol anesthesia likely contributed substantially to the lower CBF. Previous human experimental studies have demonstrated proportional reductions in CBF and cerebral metabolic rate of oxygen during propofol anesthesia, with preservation of the oxygen extraction fraction, consistent with preserved flow–metabolism coupling.^8,38^

Importantly, the marked reduction in CBF should not be interpreted as evidence of inadequate cerebral perfusion. Randomized studies have not demonstrated worse postoperative neurocognitive outcomes with propofol-based compared with volatile anesthesia.^39^ Similarly, general anesthesia during endovascular thrombectomy has not been associated with worse neurological outcomes compared with non-general anesthesia, even in patients with acutely threatened cerebral tissue.^40^ Although CBF was not directly assessed and higher recanalization rates with general anesthesia complicate interpretation of the thrombectomy trials, these findings provide no clinical evidence that the lower CBF observed during general anesthesia necessarily represents clinically relevant cerebral hypoperfusion.

The present study has several strengths. The within-subject design, with each participant studied in the awake state and under propofol-based general anesthesia before and during steady-state NE-induced MAP augmentation, minimized interindividual variability. The inclusion of patients with age and comorbidity profiles reflecting contemporary surgical populations and the use of a routine anesthetic regimen enhance the clinical relevance of the findings.^41^

An additional strength is the direct quantification of absolute CBF using PC-MRI rather than relying on surrogate measures such as transcranial Doppler-derived blood flow velocity. PC-MRI has been validated against ^15^O-H2O PET, widely regarded as the reference standard for quantitative CBF assessment.^23,42^ Furthermore, its excellent repeatability and low measurement variability make PC-MRI particularly well suited for detecting changes in CBF.^30^

Several limitations should be acknowledged. First, the study included a relatively small number of participants, although the prospectively determined sample size provided adequate power for the prespecified primary endpoint. Second, the study population consisted of neurologically intact patients undergoing elective surgery; the CBF response to MAP augmentation may not be generalizable to patients with acute cerebral pathology or impaired cerebral autoregulation. Finally, only propofol-based anesthesia was investigated, and the observed responses may differ with volatile anesthetic agents.

## Conclusion

In neurologically intact patients undergoing propofol-based general anesthesia, clinically relevant NE-induced MAP augmentation did not increase CBF. General anesthesia was associated with an approximately 50% reduction in CBF compared with the awake state, a reduction that persisted during MAP augmentation. These findings challenge the assumption that pharmacological elevation of an otherwise adequate MAP improves cerebral perfusion in this setting. While maintaining adequate arterial pressure remains essential, further MAP augmentation should not be assumed to increase CBF.

## Data Availability

The data that support the findings of this study are available from the corresponding author upon reasonable request.

## Acknowledgments

The authors thank Jadranka Plese, MSc (Department of Anesthesiology and Intensive Care, Umeå University Hospital, Umeå, Sweden), for her valuable assistance with participant recruitment and oversight of the study procedures; Jan Axelsson, PhD (Department of Diagnostics and Intervention, Umeå University, Umeå, Sweden), for assistance with MRI data management; and Peter Hällman, MD (Department of Otorhinolaryngology, Umeå University Hospital, Umeå, Sweden), for his support in facilitating the conduct of the study.

